# From many voices, one question: Community co-design of a population-based qualitative cancer research study

**DOI:** 10.1101/2024.06.24.24308902

**Authors:** Susannah K. Ayre, Elizabeth A. Johnston, Xanthia E. Bourdaniotis, Leah Zajdlewicz, Vanessa L. Beesley, Jason D. Pole, Aaron Hansen, Harry Gasper, Danica Cossio, Gemma Lock, Belinda C. Goodwin

## Abstract

**Purpose:** This study formed the development stage of a population-based survey aiming to: (i) understand the needs and experiences of people affected by cancer in Queensland, Australia and (ii) recruit a pool of participants for ongoing cancer survivorship research. The current study aimed to co-design and test a single qualitative survey question and study invitation materials to maximise acceptability of, and participation in, the survey and future research.

**Methods:** Fifty-two community members participated across 15 co-design workshops and 20 pretest interviews. During workshops, participants generated and refined ideas for an open-ended survey question and provided feedback on a study invitation letter. The use of a single, open-ended question aims to minimise participant burden while collecting rich information about needs and experiences. The research team then shortlisted the question ideas and revised study invitation materials based on workshop feedback. Next, using interviews, community members were asked to respond to a shortlisted question to test its interpretability and relevance and to review revised invitation materials. Content analysis of participant feedback was used to identify principles for designing study materials.

**Results:** Ten principles for designing qualitative survey questions were identified from participant feedback. Key principles included: *define the question timeframe and scope; provide reassurance that responses are valid and valued;* and *use simple wording*. For the study invitation materials, 11 principles were identified, including these key principles: *communicate empathy and sensitivity; facilitate reciprocal benefit;* and *include a ‘human element’.* The qualitative survey question and study invitation materials created using these principles were considered relevant and acceptable for use in a population-based survey.

**Conclusions:** Through community consultation and co-design, this study identified principles for designing qualitative data collection and invitation materials for use in cancer survivorship research. These principles can be applied by other researchers to develop study materials that are sensitive to the needs and preferences of community members.

## Introduction

In recent decades, an ageing population and low uptake of healthy lifestyle behaviours have resulted in more people being diagnosed with cancer, and improvements in cancer detection and treatment means people are living longer post-diagnosis [1,2]. While people diagnosed with cancer have unique disease trajectories, a common experience of cancer and its treatment is the widespread impact on a person’s health and wellbeing, often continuing well after treatment completion [3]. Informal caregivers (i.e., family and friends) are closely involved in supporting their loved ones to manage the impact of cancer and its treatment, often with minimal preparation for their caregiving role [4]. Consequently, caregivers can experience poor health and wellbeing due to the prolonged stress and physical demands involved [5]. In the context of increasingly resource-constrained healthcare systems, there is a growing need for community-based supportive care services that are effective in supporting the increasing number of cancer survivors and caregivers [6].

The delivery of effective supportive care services cannot be achieved without a comprehensive understanding of the needs and experiences of the cancer survivors and caregivers for whom these services are provided to. Evidence indicates that patient- and family-centred interventions result in higher satisfaction with healthcare, increased knowledge and skills for managing self-care behaviours, reduced reliance on healthcare services, and improved quality of life for both patients and caregivers [7]. Many instruments have been used in research and clinical practice to assess needs [8,9]. The Supportive Care Needs Survey-Short Form (SCNS-SF34) [10], Cancer Survivors’ Unmet Needs (CaSUN) [11], Comprehensive Needs Assessment Tool for Cancer Caregivers (CNAT-C) [12], and Supportive Care Needs Survey for partners and caregivers (SCNS-P&C) [13] are among the most widely used. However, such instruments rely on set items with pre-determined responses that may not capture the full scope of needs and do not allow respondents to express their experiences in their own words [14]. Thus, qualitative survey methods are necessary to achieve a comprehensive understanding of the needs and experiences of people affected by cancer at the population level. While qualitative research has typically been conducted with small samples, advancements in artificial intelligence and machine learning analytic techniques provide an opportunity to analyse large volumes of open-text data in a timely manner to inform service delivery [15]. To date, no suitable qualitative survey exists, providing an opportunity for community participation in co-designing materials to capture the supportive care needs and experiences of people affected by cancer.

Co-design research methods encompass various levels of consumer engagement in the research process, including participation in study conceptualisation, design, conduct, and reporting [16]. Consumer involvement in research can contribute to better study outcomes, including higher enrolment and retention rates [17]. Additionally, thorough pretesting of study materials with consumers is recommended to ensure materials are easy to understand, sensitively worded, and able to elicit meaningful data to address research aims [18].

Through active and repeated engagement with community members, this study aimed to: (i) develop and test a qualitative survey question for collecting rich information about the supportive care needs and experiences of people affected by cancer, and (ii) design study invitation materials that are relevant and acceptable to cancer survivors and their caregivers. The use of a single, open-ended question aims to minimise participant burden and maintain a broad investigation of supportive care needs, rather than the traditional itemised approach.

The materials developed in this study will be used in a new population-based study for understanding the needs and experiences of adults affected by cancer in Queensland, Australia and establishing a research-ready pool of participants to take part in ongoing cancer survivorship research. Findings from the current study will provide principles for researchers to apply when designing qualitative data collection tools and study invitation materials for research into supportive care needs and experiences, particularly in the cancer context.

## Materials and methods

This study comprised two phases of qualitative research: 1) *co-design workshops* and 2) *pretest interviews.* As outlined in **Fig 1**, both phases included community members and the research team working collaboratively to co-design a qualitative survey question and study invitation materials. Ethical approval for this study was obtained from the University of Southern Queensland Human Research Ethics Committee (ref: ETH2023-0140). This study is reported as per the Consolidated Criteria for Reporting Qualitative Research (COREQ) Checklist [19] (see **S1 Table**).

### Participants and recruitment

This study was conducted in Queensland, Australia. Queensland has almost one third of its population living outside of major cities [20], which presents several challenges to accessing cancer treatment and support due to the greater travel distances and costs involved [21]. For both the co-design workshops and pretest interviews, eligible participants were aged 18 years or older and English-speaking. Additional eligibility criteria for interview participants included a personal experience with cancer, either as a survivor or caregiver.

Digital and printed recruitment flyers were distributed via networks associated with Cancer Council Queensland or the broader research team between 11 October 2023 and 14 February 2024. To support the recruitment of priority populations, such as culturally and/or linguistically diverse (CALD) groups, the research team submitted study information to a health consumers network [22] for inclusion in their e-newsletter. As data collection advanced, recruitment was supplemented through snowball sampling, with workshop participants invited to share recruitment flyers with friends, family members, and colleagues.

Prospective participants self-enrolled into a workshop and/or interview via an online participant information and consent form administered through REDCap (hosted by the University of Southern Queensland) [23,24]. Consenting participants were then contacted by telephone and/or email to arrange a time for their participation. For the interviews, new participants were recruited alongside eligible workshop participants to obtain feedback from people with and without prior knowledge of the study.

Recruitment for the co-design workshops and pretest interviews continued until a diverse sample had been achieved and the research questions had been adequately explored, determined by the authors through concurrent data collection and analysis. Due to the large number of online registrations for the interviews, new and existing participants were purposively sampled based on their demographic characteristics, including gender, ethnicity, Indigenous status, and geographical location, as well as their experience with cancer (i.e., survivor or caregiver) to ensure that diverse perspectives were represented [25]. To acknowledge their contributions to the study, workshop and interview participants received a voucher valued at AU$100.00 (approx. 120 minutes) and AU$50.00 (approx. 60 minutes), respectively.

## Data collection

### Phase 1: Co-design workshops

Each workshop included two to four participants and was facilitated by two female researchers with undergraduate or postgraduate degrees in health science fields and training in qualitative data collection (SA, XB, and/or EJ). The facilitators had no prior relationship with the participants. At the start of each workshop, the facilitators introduced themselves, including their role in the research team and their academic background. Workshops were conducted as either online (*n*=9), in-person (*n*=1), or hybrid (i.e., online and in-person) (*n*=5) sessions using Microsoft Teams. In-person participants attended the session at one of two not-for-profit organisations, where participants were provided with the relevant materials (e.g., pen, paper). Participants attending online were asked to source these materials themselves. Workshops were audio-recorded and transcribed via Microsoft Teams. The workshops comprised several activities, with accompanying information and instructions provided on presentation slides. An overview of the workshop protocol is available in **S2 Table**.

The first activity in the workshop used the nominal group technique [26,27] to generate and refine a pool of qualitative questions that could be used in the population-based survey for understanding the supportive care needs and experiences of people affected by cancer. This technique fosters balanced participation, serving as an effective and efficient method for achieving group consensus [27]. First, participants were asked to individually brainstorm ideas for how to word the qualitative question. A preamble to the question drafted by the research team was shared with participants as a prompt for question generation (see **S2 Table** for further details). Participants were instructed that this question should be a standalone, broad, open-ended question that allows respondents to describe their needs and experiences using their own words, with the purpose of generating data that would inform service delivery at Cancer Council Queensland. Second, participants shared their questions with the group following a ‘round-robin’ process, during which a facilitator typed the questions verbatim onto the presentation slides. Third, participants collectively reflected on the proposed questions, sought clarification from one another, and adjusted wording as needed (e.g., removing potentially insensitive words). Finally, participants privately voted on their two most preferred questions using an online or paper-based poll. Votes were tallied to identify questions with the highest scores which were then presented to the group for further discussion and refinement, resulting in a total of one to five questions per group.

The second activity in the workshop involved participants providing feedback on the wording and format of an invitation letter for the population-based survey. This letter was drafted by the research team based on an example from a previous registry study. It comprised a single page of information about the research project, including its aims, instructions on how to participate, and permissions to recruit participants via the registry. During the workshops, participants were also prompted to discuss an appropriate time post-diagnosis to invite individuals to complete the survey.

Key principles endorsed by workshop participants for developing a qualitative survey question were presented to the investigator team, alongside questions that were open-ended, broad in scope, and aligned with these principles (see *Data analysis* section for methods used to identify key principles). The investigator team, comprising clinicians, researchers, and academics with expertise in cancer survivorship, supportive care, medical oncology, behavioural science, and digital health (*n*=7; see **S3 Table**), were asked to rank their five most preferred questions (1 = ‘most preferred’, 5 = ‘least preferred’) via an anonymous online survey in REDCap [23,24]. Four of the highest ranked questions were shortlisted for testing during interviews. Study invitation materials were also revised based on key principles endorsed by participants in the workshops.

### Phase 2: Pretest interviews

Online semi-structured interviews were conducted, audio-recorded, and transcribed using Microsoft Teams. Each interview was facilitated by one researcher (SA or XB). A summary of the interview protocol is available in **S4 Table**. Participants firstly reviewed the revised invitation materials and provided feedback on their readability and design. They were also asked to discuss an appropriate time for sending individuals a reminder letter to complete the survey. Participants were then randomly presented with one of the four shortlisted questions and given five uninterrupted minutes to respond to and submit their written response via the online chat function. Drawing on principles of the ‘think aloud’ method [28], participants then verbalised their thoughts, assumptions, and decisions while reading, interpreting, and responding to the question.

Scripted and spontaneous probes were used to clarify interpretation as needed. Participants were then presented with the three alternative questions and asked to nominate their preferred question based on interpretability and relevance. After 14 interviews, there was a clear consensus on the most appropriate question to include in the survey (*n*=13 votes). A further six interviews were then conducted to optimise and finalise the wording of the selected question using the ‘think aloud’ method [28] to confirm its interpretability and relevance. Following the 20 interviews, feedback on the revised study invitation materials were applied by the research team.

### Demographic survey

Age, gender, country of birth, language spoken at home, ethnicity, postcode of residential address, and personal history with cancer (including patient or caregiver status) were collected via an online survey in REDCap [23,24] at the start of each co-design workshop. For participants who did not attend a workshop, demographic data were collected through structured questions at the end of their interview.

### Data analysis

Descriptive statistics were used to summarise participant characteristics for the co-design workshops and pretest interviews. Transcripts generated via Microsoft Teams were reviewed for accuracy alongside the audio recordings. Transcripts were then analysed using content analysis to identify key principles endorsed by participants for developing a qualitative survey question and designing study invitation materials. Content analysis involves the systematic coding of text into categories based on the words and language used, centring participants’ voice in the analysis [29]. Each transcript was coded by one author (XB, SA, or EJ), with decisions regularly discussed with other authors and documented using an audit trail.

## Results

In total, 15 co-design workshops with 44 participants and 20 pretest interviews were completed (see **Fig 2**). Twelve of the 20 interview participants had also completed a co-design workshop. The characteristics of participants in the two phases of consumer consultation are summarised in **Table 1**. Both phases included representation from population subgroups, including 5-7% who identified as Aboriginal and/or Torres Strait Islander, 9-15% who used English as a second language, 20% who were born overseas, and 27-30% who lived in a rural area.

**Table 1.** Characteristics of participants in the co-design workshops (*n*=44) and pretest interviews (*n*=20) to design materials for a population-wide study on the supportive care needs and experiences of people affected by cancer.

| Characteristic | Co-design workshops<br>N (%)<sup>a</sup> | Pretest interviews<br>N (%)<sup>a</sup> |
| --- | --- | --- |
| **Age (years)** |  |  |
| Median | 43 | 39 |
| Range | 23-79 | 30-64 |
| **Gender** |  |  |
| Female | 32 (73%) | 12 (60%) |
| Male | 12 (27%) | 8 (40%) |
| **Born overseas** |  |  |
| No | 33 (75%) | 16 (80%) |
| Yes | 9 (20%) | 4 (20%) |
| Unknown | 2 (5%) | - |
| **Ethnicity** |  |  |
| Caucasian | 36 (82%) | 15 (75%) |
| Asian | 5 (11%) | 4 (20%) |
| Aboriginal and/or Torres Strait Islander | 3 (7%) | 1 (5%) |
| **Language spoken at home** |  |  |
| English only | 38 (86%) | 17 (85%) |
| Other | 4 (9%) | 3 (15%) |
| Unknown | 2 (5%) | - |
| **Geographic remoteness (ARIA)** |  |  |
| Major city | 32 (73%) | 14 (70%) |
| Regional or remote | 12 (27%) | 6 (30%) |
| **Area-level socioeconomic status (SEIFA)<sup>b</sup>** |  |  |
| High | 24 (54%) | 7 (35%) |
| Medium | 13 (30%) | 7 (35%) |
| Low | 7 (16%) | 6 (30%) |
| **Personal experience with cancer** |  |  |
| Cancer survivor | 16 (36%) | 10 (50%) |
| Cancer caregiver | 17 (39%) | 9 (45%) |
| Both | 2 (5%) | 1 (5%) |
| Neither | 9 (20%) | - |
| **Time since diagnosis** |  |  |
| <1 year | 4 (9%) | 2 (10%) |
| 1-5 years | 6 (14%) | 3 (15%) |
| >5 years | 7 (16%) | 5 (25%) |
| Unknown | 1 (2%) | 1 (5%) |
| Not applicable | 26 (59%) | 9 (45%) |
| **Duration of caregiving** |  |  |
| <1 year | 7 (16%) | 3 (15%) |
| 1-5 years | 6 (14%) | 5 (25%) |
| >5 years | 5 (11%) | 1 (5%) |
| Unknown | 1 (2%) | - |
| Not applicable | 25 (57%) | 10 (50%) |

**Abbreviations:** ARIA, Accessibility/Remoteness Index of Australia; SEIFA, Socio-Economic Indexes for Areas
<sup>a</sup> Number and percentage unless otherwise stated.
<sup>b</sup> Higher scores indicate higher relative socioeconomic advantage and lower relative socioeconomic disadvantage in general (vice versa for lower scores). High = deciles 7-10, medium = deciles 4-6, low = deciles 1-3.

### Qualitative survey question

Analysis of the 15 co-design workshop responses yielded 16 eligible questions (see **Table 2**). Additionally, 10 principles were identified from participant feedback during the workshops for developing qualitative data collection tools (see **Table 3**). These principles were to: *avoid assumptions and leading questions; define the question timeframe and scope; directly address the respondent; foster a collectivist perspective; gather experiential data from respondents for researchers to identify solutions; prompt an open-minded response; provide reassurance that responses are valid and valued; use an engaging design and accessible formatting* (e.g., large and easy to read text)*; use sensitive language;* and *use simple wording*.

**Table 2.** Eligible questions generated in co-design workshops for designing a population-wide study on the supportive care needs and experiences of people affected by cancer.

| Eligible questions<sup>a</sup> |  |
| --- | --- |
| 1 | What is the most important thing for you right now? |
| 2 | Please tell us how we can support you right now? |
| 3 | How has cancer affected your life and what do you need right now? |
| 4 | What support do you need right now based on your experience? |
| 5 | **Please tell us about your experiences since diagnosis and what would help you most right now?** |
| 6 | Tell us what we need to know right now. |
| 7 | What would you find helpful right now? |
| 8 | What does your experience look like right now? |
| 9 | What support do you need right now? |
| 10 | How can we best support you to ensure your physical, emotional, and practical needs are met right now? |
| 11 | For you to feel supported, what do we need to do right now? |
| 12 | How can we support you right now? |
| 13 | How has your life been impacted right now? |
| 14 | Please share both your positive and negative experiences since diagnosis. |
| 15 | What kind of help do you need right now? |
| 16 | Please share your experiences and challenges since your diagnosis. |

<sup>a</sup> The order of questions was randomly assigned for coinvestigator voting and listed as shown in this table. Questions in bold type were the four highest ranked questions for further testing in interviews based on preferential voting by the co-investigator team.

**Table 3.** Principles endorsed by participants during co-design workshops for developing a qualitative survey to collect information on supportive care needs and experiences of people affected by cancer in a population-based study.

| Principle | Explanation | Illustrative quotes from co-design workshops<sup>a</sup> |
| --- | --- | --- |
| **Avoid assumptions and leading questions** | Avoid making assumptions about how respondents might experience a cancer diagnosis, to ensure that those with diverse perspectives feel included, and to promote varied responses. For example, terms that imply a negative experience, such as ‘challenges’ and ‘struggles’, could limit the range of responses provided. | • **P1:** “Leaving it broader gives opportunity to include [responses like], ‘I did have a good support service, these people were really valuable’”. (**Workshop O**)<br>• **P23:** “[If asking respondents what services are lacking], you’re assuming that people think they are lacking... There could be people that are thrilled with what they are receiving... Assuming they’re lacking may not be helpful”. (**Workshop G**) |
| **Define the question timeframe and scope** | Provide guidance on what type of response would be relevant to the research aims. For example, include a timeframe for the question (e.g., ‘since diagnosis’) and prompts to define its scope. These prompts should encourage detail and allow for variation in respondents’ answers (e.g., ‘Be as specific as possible’ and ‘You might like to consider your personal experiences with employment, transport, grief...’). Including question prompts helps to minimise burden and manage participant expectations of the research outcomes. | • **P15:** “I need some specificity [about the timeframe]... Are you talking about [my needs and experiences] through treatment or are you talking about now, or are you talking about after? That’s the specificity I would need to answer that question”. (**Workshop J**)<br>• **P35:** “The only thing I would suggest is prompting [people to think about] different types of experiences – financial, social, emotional, relational. I think a lot of people think, ‘I’m having chemo’ and ‘I’m having surgery’ and ‘I’m feeling sick’, but there is also the isolation... not being with your friends and family. That ties into mental health and wellness. Then [there are the] financial and practical [impacts] too”. (**Workshop C**)<br>• **P38:** “Come up with a list of what Cancer Council Queensland can do... So, the person that is confronting this thing [thinks], ‘Well alright, I can get financial counselling from you or help getting to the hospital’. ...I think it would help to provide a bit of an overview of the general types of services you might be referring to [in this question]”. (**Workshop B**) |
| **Directly address the respondent** | Use second-person language to directly address and engage the respondent. | • **P26:** “I like the ‘you’ and ‘we’ because it is more direct. It engages you and you feel that you are being addressed”. (**Workshop F**) |
| **Foster a collectivist perspective** | Frame the question so that it encompasses the broader community, rather than the individual alone. Framing the question as such may encourage answers from respondents who prioritise the experiences of others before their own or may not feel comfortable expressing their own need for support. | • **P23:** “[The question] could be more generally about... how can we make what we do even better for *everybody*? Because culturally that is how a lot of people think... They tend to identify the general experience for everyone rather than the individual”. **(Workshop G)**<br>• **P26:** “[If asked about what support I need], back then, I would have said nothing. I would have been like, ‘No, I’m doing this myself. I can handle it’. Whereas if the wording... was like, ‘How can we support people with cancer?’, I’d probably be more likely to answer it... It would still really be about my experience, but it would be looking out for other people instead of myself”. **(Workshop F)**<br>• **P42:** “From my partner who was volunteering [in a wigs service], she mentioned that ladies would go there, but they really weren’t worried about having a wig. They were getting the wig to not scare their grandkids... Some things they do is purely for [their family]... So, [I like] ‘What would make life easier for you and those you care for?’” **(Workshop A)** |
| **Gather experiential data from respondents for researchers to identify solutions** | Invite respondents to share their personal experiences, rather than only focusing on their needs or asking them to provide a solution to address their needs. Participants felt that solutions for addressing supportive care needs are best devised with researcher input. | • **P15:** “I feel like it’s difficult for people not really involved in [research] to come up with their own thoughts on what services [they need]. They don’t know what services are out there... ‘What services do I want? Well, what’s something that even exists?’ ...I feel like if you are not someone who is involved in this area, it might be hard for you to come up with these ideas on your own”. **(Workshop J)**<br>• **P30:** “I know when I was jotting down [ideas for] the wording of the question, I started with asking people to share their experiences, rather than detailing what support services they want. [The latter] seems a bit direct and too specific”. **(Workshop E)**<br>• **P35:** “You often don’t know what you need... because your brain is in survival mode to get through each day. I like the idea of sharing your experience and then the people... behind the research can, with a trained eye, can look at what are people struggling with? What has their experience been? What are the gaps that we can see?” **(Workshop C)** |
| **Prompt an open-minded response** | Encourage respondents to think beyond the scope of support that is currently available. | • **P26:** “If you put ‘hope’ or ‘wish’ [in the question], it might help [respondents] expand [their answer]. It’d have them [think], ‘Oh, I wish someone could...’”. (**Workshop F**)<br>• **P42:** “I feel like ‘In a perfect world...’ helps people think about not just what’s on offer, but [what they would want] if they could have anything. They will actually share anything they want. Rather than saying, ‘Okay, what support do I have?’ [and] writing that down”. (**Workshop N**) |
| **Provide reassurance that responses are valid and valued** | Include general prompts to provide reassurance that all answers are valid and valued. For example, ‘There are no wrong answers,’ and ‘You can share as much or as little as you feel comfortable with’. | • **P29:** “If you want people to give a lot of detail, you need to be explicit about that. For example, ‘In as much detail as you feel alright to share...’. (**Workshop E**)<br>• **P30:** “Put in the question that, ‘It is as much as you feel comfortable to share’. So, [respondents] don’t feel like their response won’t be included if they don’t provide a lot of detail”. (**Workshop E**) |
| **Use an engaging design and accessible formatting** | Use formatting that encourages respondents to answer. For example, use large font, include the survey on one page, and do not use a character or word limit in the online survey. | • **P2:** “I think [the survey] should be on one page. From my own experience, what you read in the first paragraph, or the first couple of paragraphs, is what sells you... You don’t want to read a whole lot of stuff”. (**Workshop O**)<br>• **P5:** “If there’s older people with... vision impairments, is the text big enough? Make sure it’s accessible”. (**Workshop N**)<br>• **P28:** “Sometimes when people get on a roll telling their story, they need more [space]... If you limit [their answer] to 1,000 characters, people will think they are running out of character space, [and think], ‘Let me go back to see what I can eliminate to get my story in’”. (**Workshop E**) |
| **Use sensitive language** | Avoid potentially insensitive terms, such as ‘live well,’ ‘journey,’ and ‘fighting’ as these terms imply a degree of choice, control, or contentment in a cancer diagnosis. | • **P6:** “Get rid of ‘fighting’. A lot of [cancer patients] don’t like it... Sometimes people don’t have a fight. Sometimes they do and they feel survival guilt... It is one of those triggering words”. (**Workshop N**)<br>• **P28:** “I don’t like the word ‘journey’. It implies that cancer is a holiday and nothing about cancer is a holiday”. (**Workshop F**)<br>• **P37:** “[Remove] ‘live well’ [from the preamble], especially for people that might have received a terminal diagnosis. Unfortunately, they are not going to live well. It is a hard slog through that”. (**Workshop B**) |
| **Use simple** | Use simple and short sentences. Avoid | • **P23:** “Sentences such as, ‘What does this [support] *look like* to you?’ could be too |
| **wording** | phrases that could be misinterpreted by people for whom English is a second language. | vague... People might think you are asking for a physical description [of what the services look like]. ...If they are older, or [use English as a] second language, that sentence wouldn't make sense". (**Workshop G**)<br>• **P26:** "[Removing the term 'services' makes the question] more digestible, and not as intimidating and overwhelming for someone with lower literacy. It's the word 'services' that intellectualises the question". (**Workshop F**)<br>• **P42:** "Keep the question as simple as possible so that more people can understand the question and therefore answer". (**Workshop A**) |

273 Participants have been designated a code to maintain their anonymity.

A flowchart showing idea generation and shortlisting for the qualitative survey question is presented in **Fig 3**. During the workshops, participants generated a total of 173 questions with 42 questions selected through participant voting. Of these 42 questions, 14 (33%) did not meet pre-defined criteria (i.e., not open-ended, not broad in scope) and 1 question was a duplicate, warranting their exclusion. An additional 11 (26%) questions were excluded for not aligning with the principles endorsed by workshop participants. Although participants advocated for including a timeframe within the question, a limited number of questions aligned with this principle. Rather than excluding these questions, a timeframe was added as needed (e.g., ‘right now’ was added to Question 1; see **Table 2**). Similarly, few questions complied with the principles of prompting an open-minded response (e.g., ‘in an ideal world’) and fostering a collectivist perspective (e.g., ‘you or those you care for’). Questions that did not align with these principles were not excluded as the population-based study aimed to capture actual needs and experiences from the perspective of individual respondents.

From the 16 eligible questions, preferential voting by the investigator team resulted in a shortlist of 4 questions. In line with principles endorsed by workshop participants, team members were in favour of including prompts alongside the question, with the most popular prompts being: ‘Share as much detail as you would like to’ (*n*=5, 71%) and ‘For example, you might like to consider your practical, emotional, psychological, financial, relational, and cultural needs’ (*n*=5, 71%). **Table 4** summarises participants’ interpretations of and feedback on these questions and prompts in the pretest interviews. Participants’ responses to the shortlisted questions are provided in **S5 Table** for comparison.

**Table 4.**
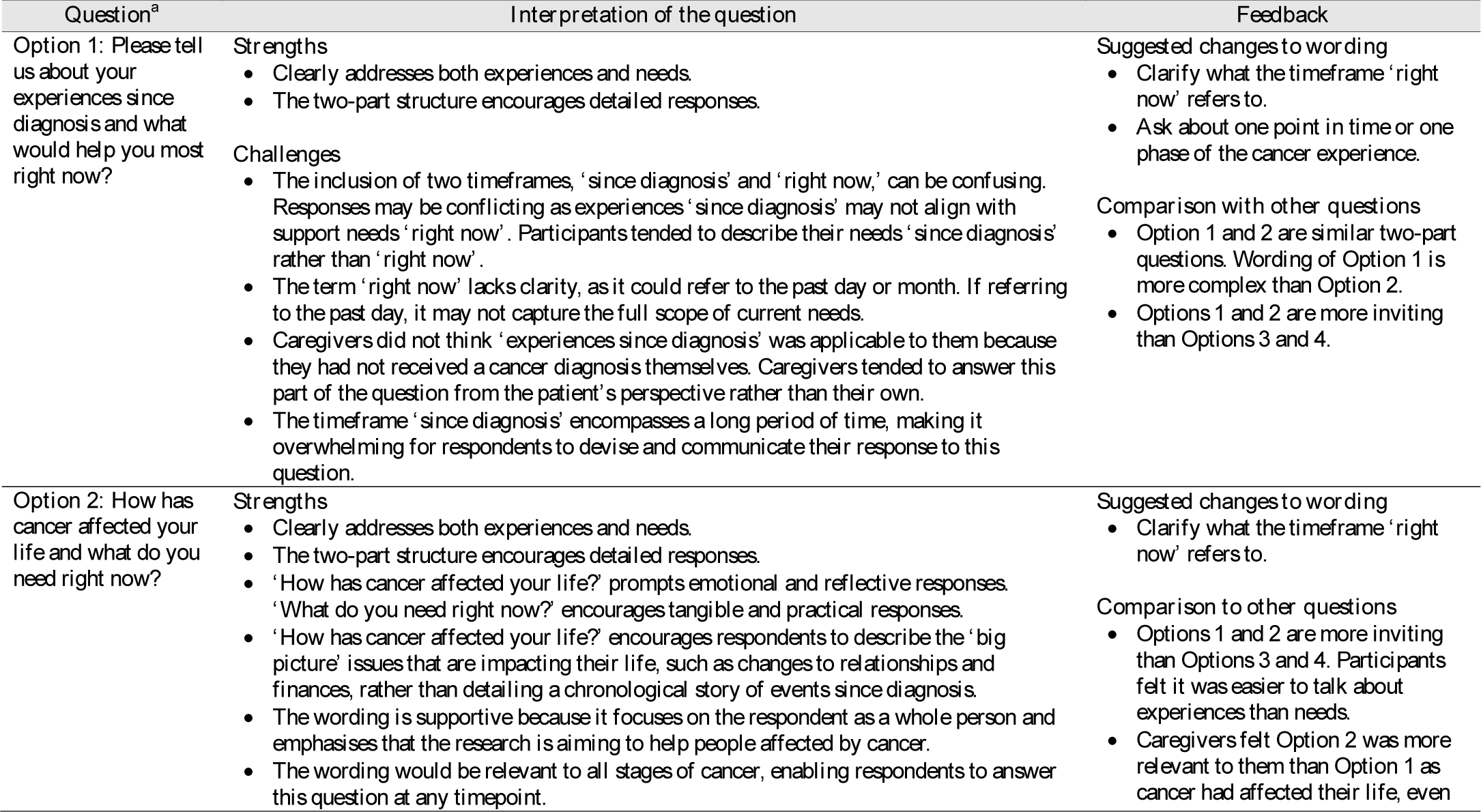
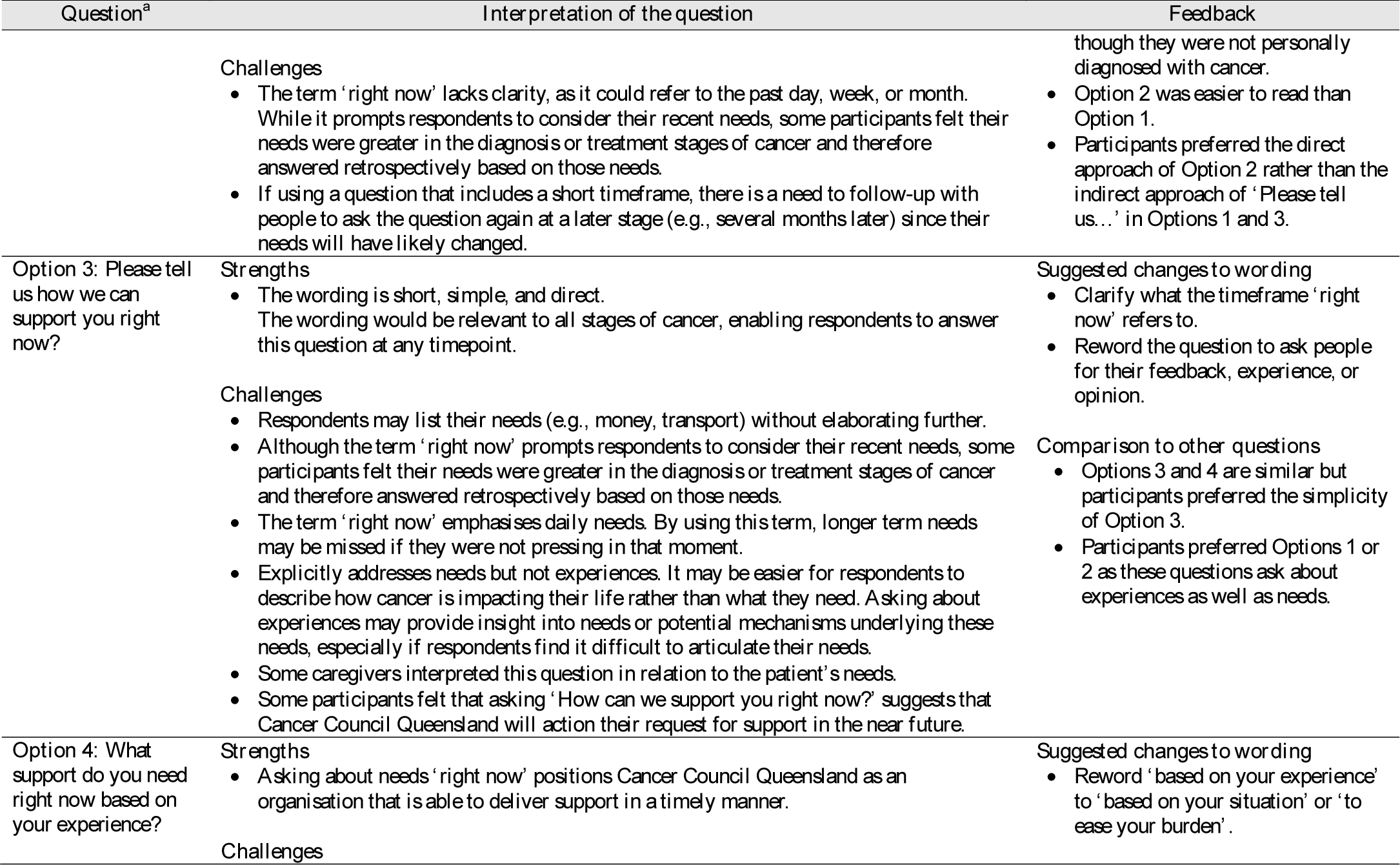
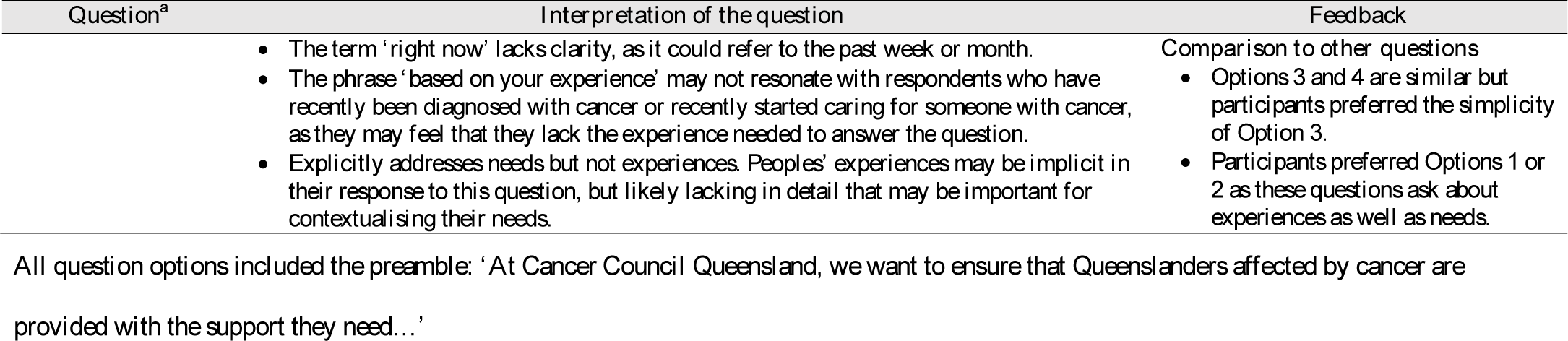
Feedback from participants in the pretest interviews regarding the four shortlisted questions for a population-wide study on the pportive care needs and experiences of people affected by cancer.^a^.

Of the four shortlisted questions, interview participants preferred the two-part questions that explicitly asked about needs *and* experiences (i.e., Options 1 and 2) (see **Table 4**). Participants thought this question structure would generate richer information, particularly from respondents who find it difficult to articulate their needs. Of these two options, most participants (*n*=13/14) endorsed Option 2 as it was easier to read and interpret, applicable to both cancer survivors and their caregivers, and relevant to any timepoint post-diagnosis.

Participants recommended clarifying the timeframe that ‘right now’ referred to. For the question prompts, it was suggested to substitute complex words (i.e., replace ‘relational’ with ‘social’), remove words with similar meanings (i.e., remove ‘psychological’ and retain ‘emotional’), and broaden the scope (i.e., add ‘physical’ and ‘spiritual’). Option 2 was revised accordingly, with the wording of this question also changed to present tense to provide further clarification of the timeframe. These revisions resulted in the final question:

*Thinking about the past month, how is cancer affecting your life and what do you need? Share as much detail as you would like to. For example, you might like to consider your physical, emotional, practical, financial, social, cultural, and spiritual needs, or any other needs you might have*.

In the final six pretest interviews, this question demonstrated capacity to elicit relevant, detailed data on the unmet supportive care needs and experiences of people affected by cancer (see **S5 Table**).

### Study invitation materials

Ten principles for designing study invitation materials were identified from participant feedback during the co-design workshops and validated during the pretest interviews. These principles are presented in **Table 5** alongside an explanation and participant quotes. These principles were to: *communicate empathy and sensitivity; consider appropriate timing; convey credibility and legitimacy; facilitate reciprocal benefit; include a ‘human element’; increase accessibility and ease of participation; optimise readability; promote inclusivity; provide reassurance around privacy; motivate and incentivise participation;* and *support informed decisions*. An eleventh principle, *promote inclusivity*, emerged from the interview data only (see **Table 5**). **Table 5** also provides examples of how each principle was applied in this study to develop invitation materials for the proposed population-based survey. For example, following the co-design workshops, a flyer was created to accompany the letter, which included a personal quote from a cancer survivor emphasising the value of the research for people impacted by cancer.

**Table 5.**
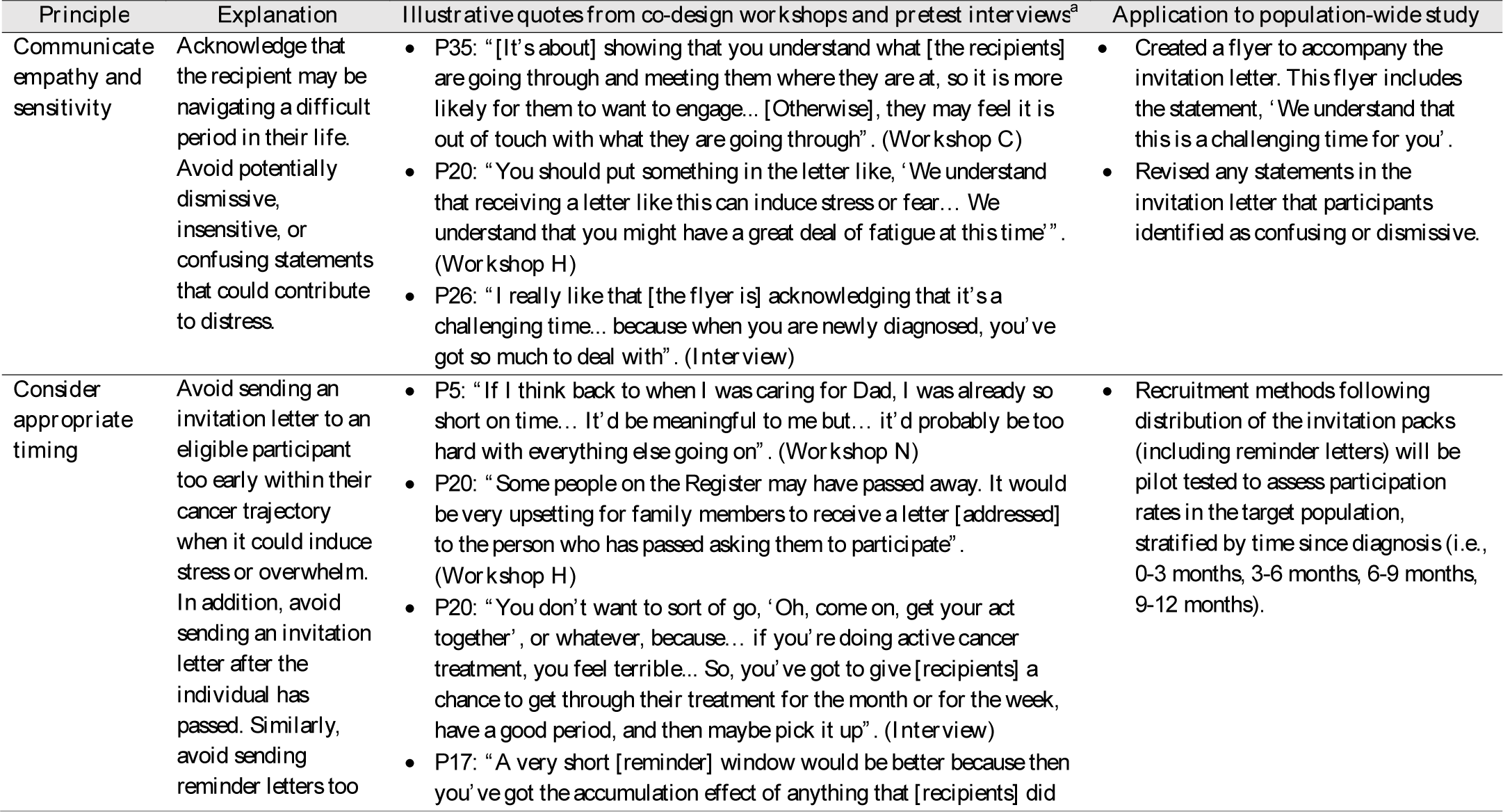
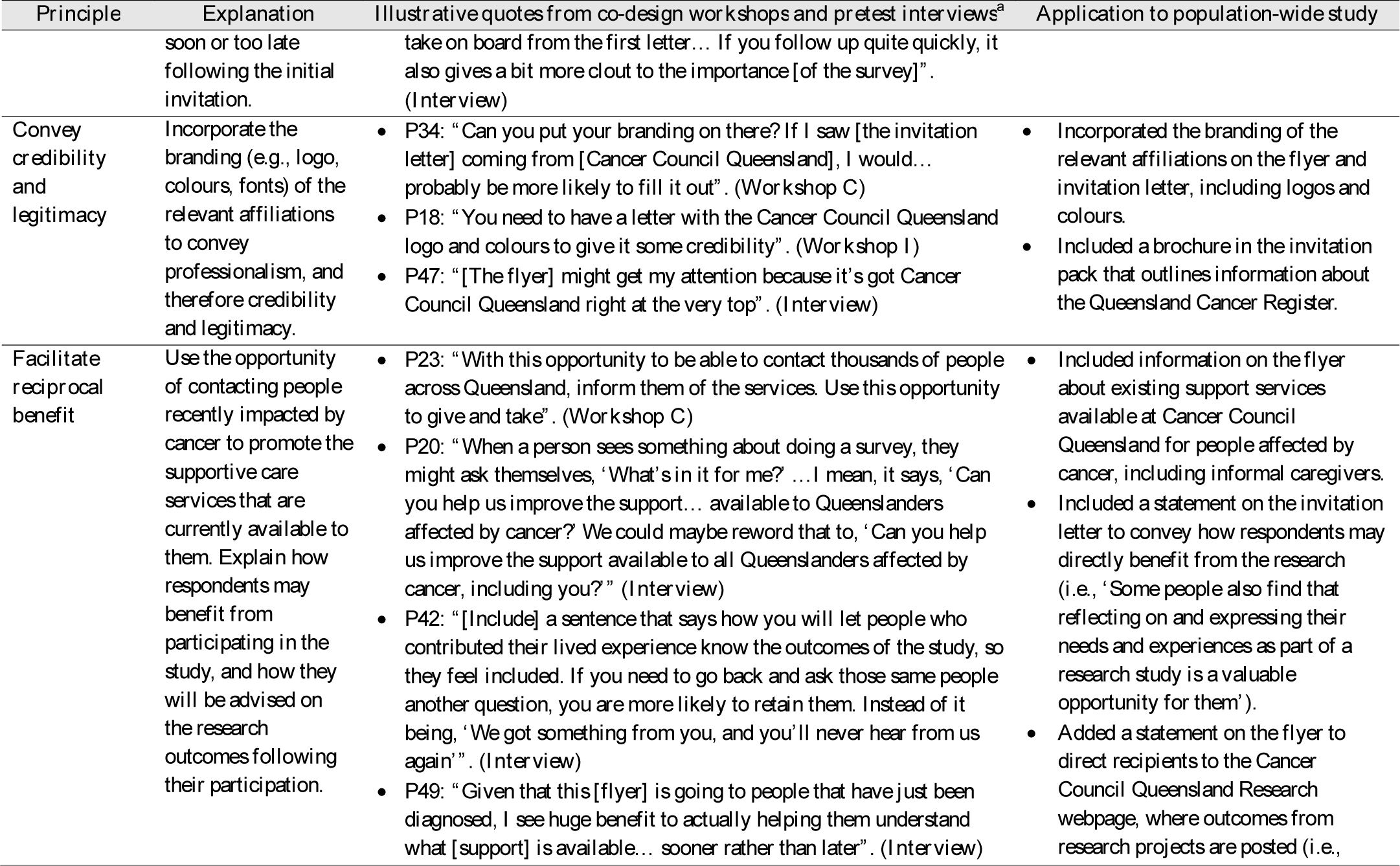
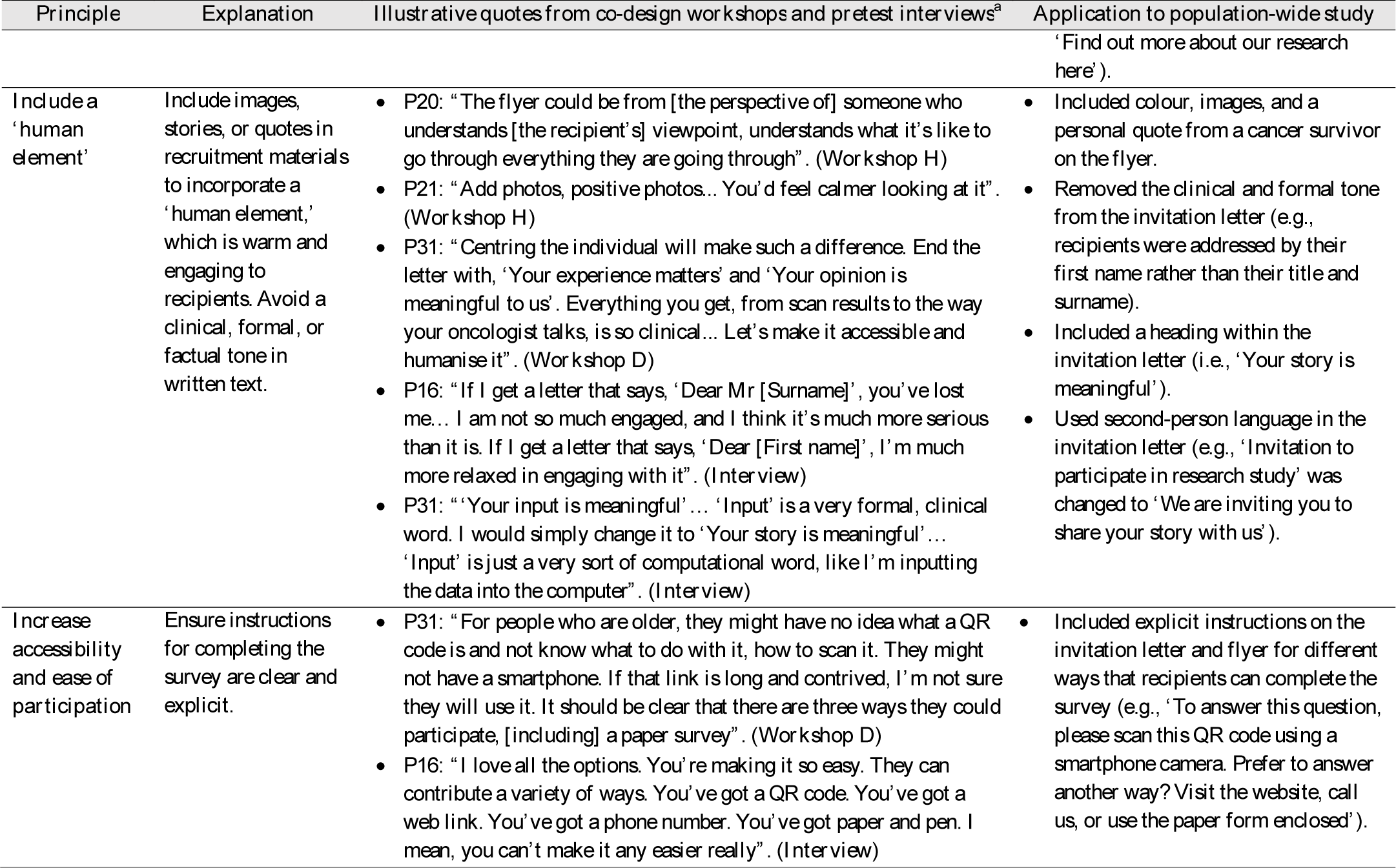
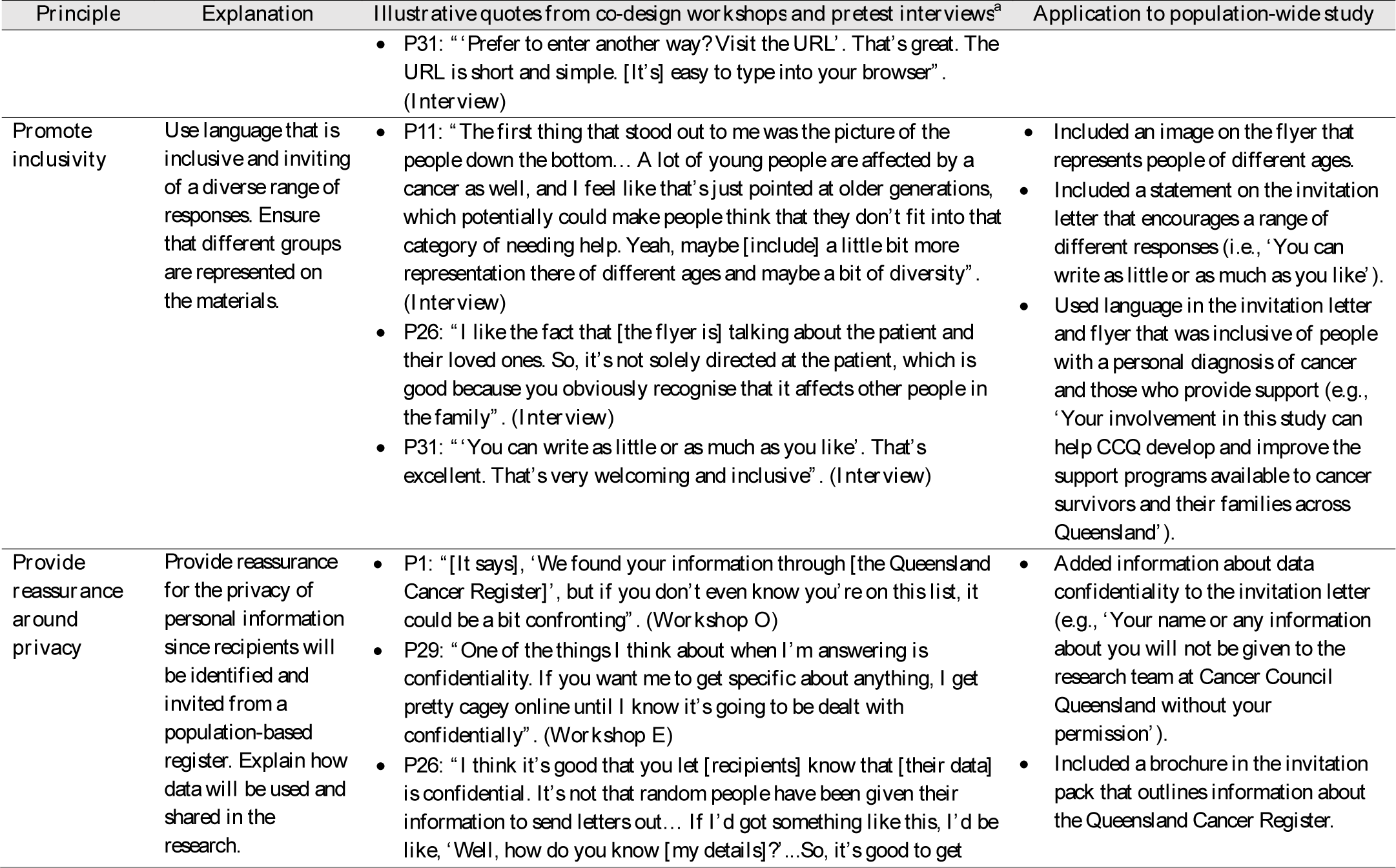
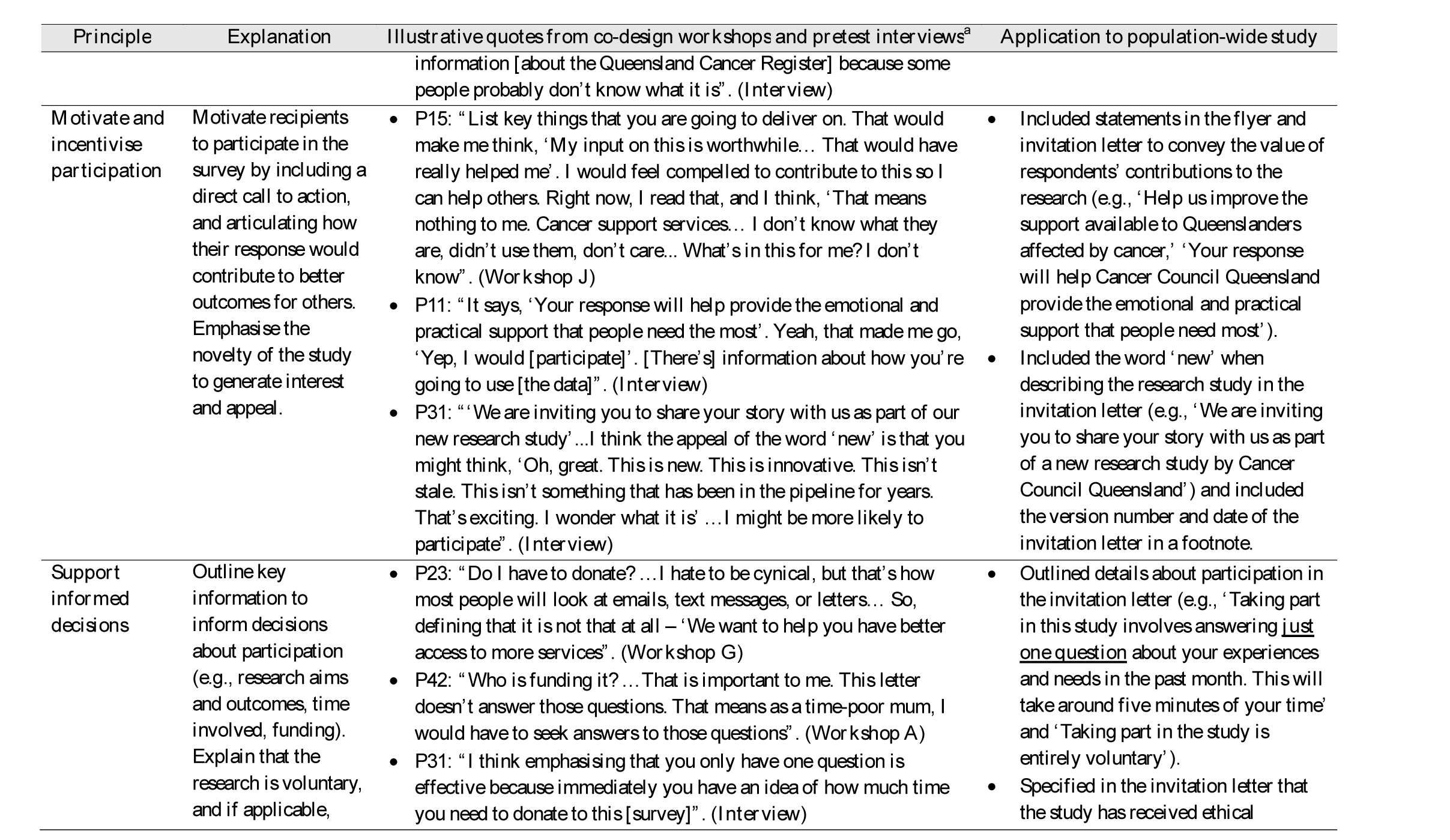
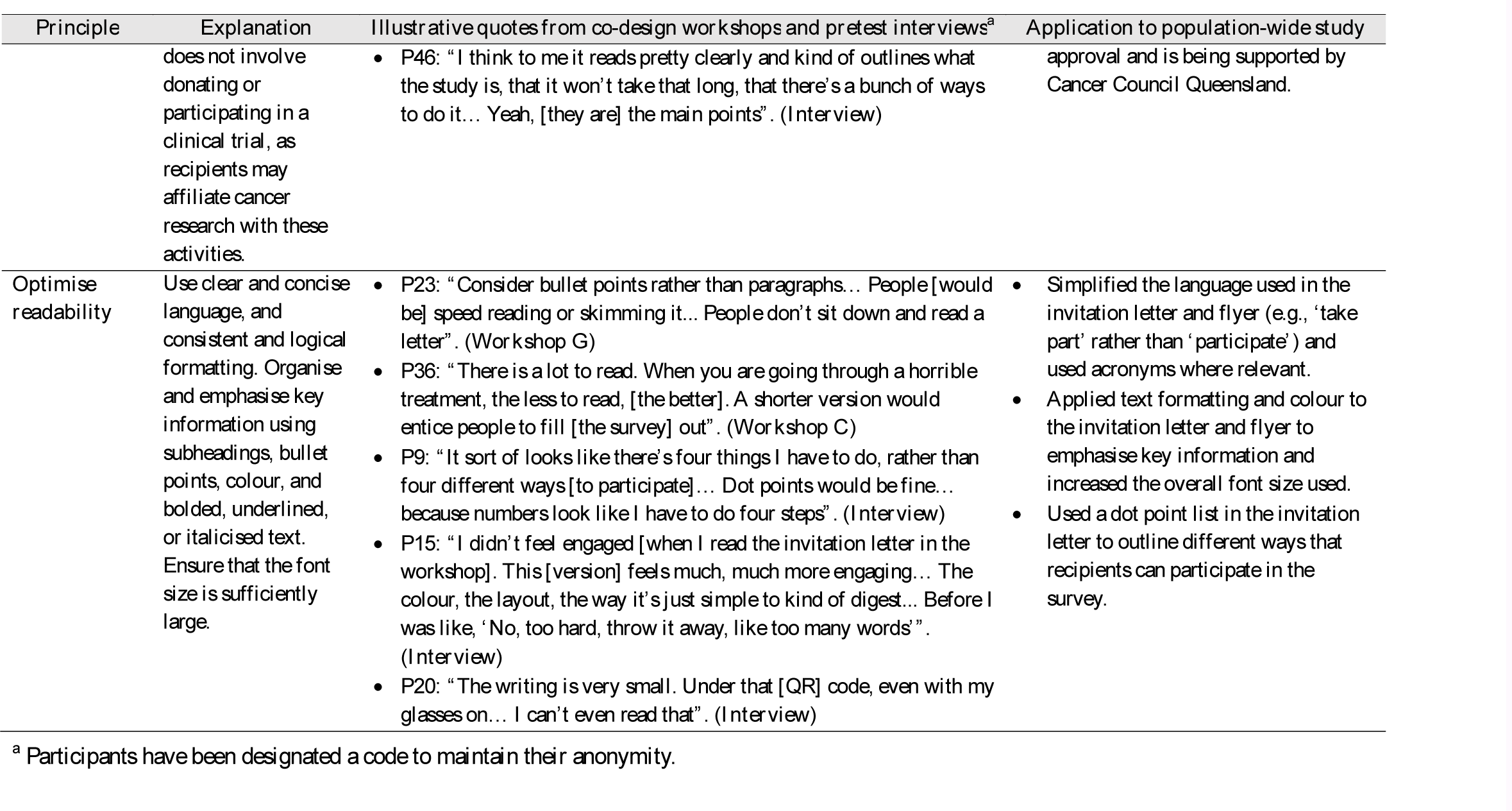
Principles for designing study invitation materials endorsed by participants during the co-design workshops and pretest interviews for co-igning a population-wide study on the supportive care needs and experiences of people affected by cancer.

The revised invitation materials presented to participants in the 20 pretest interviews were highly accepted, with 19 (95%) participants indicating they would likely respond to the qualitative survey question. The one participant who indicated they would not complete the survey was reluctant to scan a quick response (QR) code. Therefore, instructions on alternative methods for completing the survey were emphasised on the invitation materials in the final revision stage, alongside several other minor changes, such as increasing the font size and simplifying the language used.

## Discussion

This qualitative study used co-design methods to develop and test study materials for capturing the supportive care needs and experiences of cancer survivors and their caregivers in a future population-based survey. Principles for designing qualitative data collection tools and study invitation materials were identified in workshops with community members and validated in pretest interviews. These principles can be used more broadly by health and survivorship researchers to design study materials that align with community preferences.

To date, studies examining strategies to optimise participation in cancer research have focused primarily on methods of advertisement (e.g., social media, text messages), incentives, eligibility criteria, and outcome measures [30,31]. However, few studies have investigated community preferences for invitation materials. A conceptual model developed by Chhatre and colleagues [32] describes strategies for recruiting cancer patients into clinical trials using a patient-centred approach. Recommended strategies mapped onto four concepts, including trust, communication, expectations, and attitudes, and ranged from protecting patient health information to emphasising the altruistic value of research involvement [32]. These findings align with principles endorsed by community members in the current study, which supports the applicability of these principles across clinical and observational research settings.

To the authors’ knowledge, few studies have reported on the development and testing of qualitative surveys for health research, despite the importance of question wording in survey-based studies [33]. Principles identified in the current study both validate and expand on those previously identified in best practice guidelines for qualitative research [33–35]. For example, similar to consumers in this study, Braun and colleagues [33] recommend that survey questions are short and unambiguous, void of assumptions, and include examples to guide the scope of responses. Similar to other widely used surveys [10–12], a timeframe was also added to the question (i.e., the past month) to minimise participant burden when responding, enable repeated data collection over time, and allow comparability of responses by time since diagnosis. The current study also identified additional principles for designing a qualitative survey. For example, leveraging the expertise of respondents by asking them to share their experiences, rather than just their needs, enables researchers to identify trends and gaps in care at the population-level. Considering that the principles in this study were endorsed by a diverse sample of cancer survivors and caregivers, the principles identified may also be applicable to studies investigating the supportive care needs and experiences of people affected by other health conditions.

The single, open-ended survey question designed in the current study was developed to collect rich, detailed information on the supportive care needs and experiences of people affected by cancer for the purpose of informing service delivery. Current literature on supportive care needs is largely based on data derived from quantitative measures. These measures are confined to specific supportive care domains, with needs in the cognitive, spiritual, sexual, or financial domains often overlooked or excluded from shorter versions of tools [14,36]. In contrast, the question developed in this study asks about supportive care needs in the context of participants’ experiences (i.e., how cancer is affecting their life). Based on participant feedback in the workshops, the question includes a prompt that lists various domains of potential need. When this prompt was tested in interviews, its inclusion did not appear to constrain or direct participant responses. Therefore, the question provides an opportunity for respondents to share their experiences and to define what is important to them. The final question demonstrated applicability and acceptability among a diverse sample of cancer survivors and caregivers. Thus, the question presents a novel method for assessing the supportive care needs and experiences of people affected by cancer.

In the current study, there were some divergent views among community members regarding how to design a qualitative survey question. For example, some participants suggested asking about supportive care needs in an open-minded manner, using phrases such as ‘in an *ideal* world’ and ‘what support do you *wish* you had’. However, this principle conflicted with other participants’ suggestions to define the scope of the question by providing realistic examples of what supportive care services may be provided, to manage expectations of the research outcomes. Similarly, some participants noted that using terms like ‘ideal’ could be insensitive as they may imply a sense of choice or control in a person’s cancer trajectory. Therefore, the principle of asking about needs in an open-minded manner was not applied in the current study but may be suitable to other populations or contexts depending on the research aims and scope.

### Strengths and limitations

This study used an iterative qualitative design, facilitating active and repeated engagement of community members in developing and testing study materials. Including a subset of participants in both phases of consumer consultation enabled the principles identified in workshops to be validated through member checking in interviews [25]. The use of two different consultation methods was another key strength; the workshops fostered collaborative group discussion which supported idea generation, while the interviews provided a platform for in-depth exploration of individual perspectives and experiences [25]. Finally, the study included a diverse sample, involving people who were born overseas, used English as a second language, identified as Aboriginal and/or Torres Strait Islander, and lived in a rural area.

The main limitation of this study is the potential for self-selection bias. The consumer consultations relied on individuals agreeing to participate in a research study and being able to speak English. Given that an estimated 900,000 people in Australia have low proficiency in spoken English [37], and that CALD groups experience poorer health outcomes compared to the general population [38], future research should work with non-English speaking people to design study materials that facilitate their participation in cancer research [39].

## Conclusions

Through active and repeated consultation with community members, this study identified principles for designing qualitative data collection tools and study invitation materials for use in cancer survivorship research. These principles were used to design and test an open-ended survey question and study invitation materials for use in a population-based study of the supportive care needs and experiences of cancer survivors and their caregivers. These principles can also be used by other researchers to optimise community participation in their qualitative research and to inform support service providers about the needs and experiences of consumers.

## Data Availability

All data produced in the present study are available upon reasonable request to the authors.

## Acknowledgements

The authors thank all individuals who participated in a workshop and/or interview.

## Supporting information

**S1 Table:** Completed Consolidated Criteria for Reporting Qualitative Research (COREQ) checklist.

**S2 Table:** Overview of protocol for co-design workshops.

**S3 Table:** Expertise of the co-investigator research team who voted on the questions generated from the co-design workshops for inclusion in the pretest interviews.

**S4 Table:** Overview of protocol for pretest interviews.

**S5 Table:** Responses to the shortlisted and final question in the pretest interviews.

---

**Fig 1.** Overview of study procedures to co-design materials for a population-wide study on the supportive care needs and experiences of people affected by cancer.

**Fig 2.** Flowchart of participant recruitment and selection for the co-design workshops and pretest interviews to design materials for a population-wide study on the supportive care needs and experiences of people affected by cancer.

^a^Invalid responses were identified based on a combination of factors (e.g., duplicated IP addresses with different names, invalid postcodes or phone numbers, replicated responses in a short period of time, unusual completion times). Where necessary, responses flagged as potentially invalid were investigated further through phone and/or email contact.

^b^Individuals were purposively selected to achieve maximum variation in the age, gender, ethnicity, and geographic location of participants.

^c^12 of the 20 participants in the pretest interviews also participated in a co-design workshop.

**Fig 3.** Flowchart of idea generation and shortlisting to develop a single, open-ended question for a population-wide study on the supportive care needs and experiences of people affected by cancer.

^a^i.e., Terms such as ‘quality of life’ and ‘daily living’.

^b^i.e., Terms such as ‘cancer journey’.

^c^i.e., Questions that imply a negative experience, such as ‘challenging’.

^d^i.e., Questions such as ‘What would make life easier for you?’

## Notes

### Competing Interest Statement

The authors have declared no competing interest.

### Funding Statement

This study was funded by Cancer Council Queensland.

### Author Declarations

Ethical approval for this study was obtained from the University of Southern Queensland Human Research Ethics Committee (ref: ETH2023-0140).

