## Supplementary Material for "From many voices, one question: Community co-design of a population-based qualitative cancer research study"

### **S1 Table.** Completed Consolidated Criteria for Reporting Qualitative Research (COREQ) checklist.

| Topic | Item | Guide Questions/Description | Pages |
| --- | --- | --- | --- |
| Domain 1: Research team and reflexivity | | | |
| *Personal characteristics* | | | |
| Interviewer/facilitator | 1 | Which author/s conducted the interview or focus group? | 7 |
| Credentials | 2 | What were the researcher’s credentials? (e.g., PhD, MD) | 7 |
| Occupation | 3 | What was their occupation at the time of the study? | 7 |
| Gender | 4 | Was the researcher male or female? | 7 |
| Experience and training | 5 | What experience or training did the researcher have? | 7 |
| *Relationship with participants* | | | |
| Relationship established | 6 | Was a relationship established prior to study commencement? | 7-8 |
| Participant knowledge of the interviewer | 7 | What did the participants know about the researcher? (e.g., personal goals, reasons for doing the research) | 7-8 |
| Interviewer characteristics | 8 | What characteristics were reported about the interviewer/facilitator? (e.g., biases, assumptions, interests in the research topic) | 7-8 |
| Domain 2: Study design | | | |
| *Theoretical framework* | | | |
| Methodological orientation and theory | 9 | What methodological orientation was stated to underpin the study? (e.g., grounded theory, discourse analysis, ethnography, phenomenology, content analysis) | 10 |
| *Participant selection* | | | |
| Sampling | 10 | How were participants selected? (e.g., purposive, convenience, consecutive, snowball) | 6-7 |
| Method of approach | 11 | How were participants approached? (e.g., face-to-face, telephone, mail, email) | 6-7 |
| Sample size | 12 | How many participants were in the study? | 11, Fig 2 |
| Non-participation | 13 | How many people refused to participate or dropped out? What were the reasons? | Fig 2 |
| *Setting* | | | |
| Setting of data collection | 14 | Where was the data collected? (e.g., home, clinic, workplace) | 8-9 |
| Presence of non-participants | 15 | Was anyone else present besides the participants and researchers? | NA |
| Description of sample | 16 | What are the important characteristics of the sample? (e.g., demographic data, dates) | 11, Table 1 |
| *Data collection* | | | |
| Interview guide | 17 | Were questions, prompts, guides provided by the authors? Was it pilot tested? | S2 Table, S4 Table; NA |
| Repeat interviews | 18 | Were repeat interviews carried out? If yes, how many? | NA |
| Audio/visual recording | 19 | Did the research use audio or visual recording to collect the data? | 8, 9 |
| Field notes | 20 | Were field notes made during and/or after the interview or focus group? | NA |
| Duration | 21 | What was the duration of the interviews or focus groups? | 7 |
| Data saturation | 22 | Was data saturation discussed? | 7 |
| Transcripts returned | 23 | Were transcripts returned to participants for comment and/or correction? | NA |
| Domain 3: Analysis and findings | | | |
| *Data analysis* | | | |
| Number of data coders | 24 | How many data coders coded the data? | 11 |
| Description of the coding tree | 25 | Did authors provide a description of the coding tree? | NA |
| Derivation of themes | 26 | Were themes identified in advance or derived from the data? | 10-11 |
| Software | 27 | What software, if applicable, was used to manage the data? | NA |
| Participant checking | 28 | Did participants provide feedback on the findings? | 34, Fig 1 |
| *Reporting* | | | |
| Quotations presented | 29 | Were participant quotations presented to illustrate the themes/findings? Was each quotation identified? (e.g., participant number) | Tables 3 and 5 |
| Data and findings consistent | 30 | Was there consistency between the data presented and the findings? | Tables 3 and 5 |
| Clarity of major themes | 31 | Were major themes clearly presented in the findings? | 12, 23-24, Tables 3 and 5 |
| Clarity of minor themes | 32 | Is there a description of diverse cases or discussion of minor themes? | NA |

### **S2 Table.** Overview of protocol for co-design workshops.

| Protocol | |
| --- | --- |
| Equipment | |
| *In-person workshops:*   - Food and drink catering - Name tags - Pens, paper, and sticky notes - Printed demographic surveys - Printed and laminated prompt cards | - Printed recruitment letters - Laptop and screen with presentation slides - Dictaphone   ***Online workshops:***   - Microsoft Teams |
| Set up/preparation | |
| - Ask participants to write their name on a name tag [in-person only] - Ask participants to complete the demographic survey [send link to online participants] | |
| Introduction | |
| Welcome participants | |
| Introduce the research team | |
| Provide an overview of the workshop | |
| - Administrative tasks [start audio-recording with permission from participants] - Guidelines and expectations | |
| Allow participants to introduce themselves | |
| Provide overview of the vision and mission of Cancer Council Queensland (CCQ) | |
| Introduce UNIQUE | |
| - Background and aims - Recruitment - Data collection | |
| Outline purpose of the workshop | |
| Activity 1: Survey question | |
| Introduce activity | |
| Explain criteria for the survey question | |
| - Provide examples on presentation slides - Provide preamble to question on presentation slides  – i.e., *We want to ensure that all Queenslanders affected by cancer are provided with the support they need to live well. As a person affected by cancer in Queensland…* | |
| Ask participants to individually write a list of survey question ideas (10 minutes) | |
| - Provide pen and paper [in-person only] - Provide prompt cards [type prompts into chat for online participants]  – e.g., *What do you wish you were asked at the time of diagnosis? Imagine you or a loved one are facing a cancer diagnosis. What question would you want to answer to express your needs and experiences?* | |
| Invite participants to share their survey question ideas with the group following a ‘round-robin’ process | |
| - Type questions verbatim onto the presentation slides | |
| Facilitate a group discussion using the survey question ideas as prompts | |
| – e.g., *What was your thinking behind this question?* | |
| Ask participants to privately vote for their two most preferred survey question ideas | |
| - Provide pen and sticky notes [in-person only] - Create poll in Microsoft Teams [online only] | |
| 5-minute break | |
| - Tally the votes - Type the 2-3 highest ranked survey question ideas onto the presentation slides | |
| Facilitate a group discussion using the survey question ideas as prompts | |
| – e.g., *Should we include additional instructions or prompts with this question?* | |
| Activity 2: Recruitment material | |
| Introduce activity | |
| Provide overview of the recruitment methods | |
| Ask participants to read a draft version of the invitation letter   - Display a digital version of the letter on the presentation slides - Provide paper version of the letter [in-person only] | |
| Facilitate a group discussion using the invitation letter as a prompt | |
| – e.g., *What do you think about the wording of this letter?* | |
| Ask participants for their opinion on the timing for sending this letter | |
| – e.g., *In your opinion, how early can we invite people to take part in the survey following their diagnosis?* | |
| Conclusion | |
| Invite participants to share additional thoughts or feedback | |
| Invite participants to register for the second phase of the study (pretest interviews) | |
| - Provide study flyer [email digital copy to online participants] | |
| Thank participants for their contributions | |

### **S3 Table.** Expertise of the co-investigator research team who voted on the questions generated from the co-design workshops for inclusion in the pretest interviews.

| Position | Organisation type | Area of expertise | Years working in oncology |  | Focus of work |
| --- | --- | --- | --- | --- | --- |
| Professor | University | Digital health | 16 |  | AI-assisted technology, childhood cancer |
| Associate Professor | Medical research institute | Supportive care and behavioural science | 20 |  | Supportive care self-reported outcome and intervention studies, patients and caregivers |
| Associate Professor | Not-for-profit | Behavioural science and survivorship | 8 |  | Health psychology, population-level behaviours and interventions |
| Postdoctoral researcher | Not-for-profit | Survivorship | 5 |  | Rural cancer survivors and caregivers, health behaviours, support seeking |
| Director of Medical Oncology | Rural-based hospital | Medical oncology | 10 |  | Upper gastrointestinal, pancreato-biliary, central nervous system, melanoma, breast, lung, colorectal, prostate cancers |
| Deputy Director, Division of Cancer Services | Metropolitan-based hospital | Medical oncology | 14 |  | Head and neck, genitourinary cancers |
| Executive | Not-for-profit | Supportive care and service delivery | 11 |  | State-wide operations and service delivery |
| Manager | Population-based registry | Population clinical data | 19 |  | Data management and quality |

^a^ Of the 8 research team members listed, a total of 7 responded to the online anonymous survey.

### **S4 Table.** Overview of protocol for pretest interviews.

Protocol

Equipment

- Laptop and screen with presentation slides
- Microsoft Teams

Introduction

Welcome participant

Introduce self

Provide an overview of the interview

- Administrative tasks [start audio-recording with permission from participant]

Provide overview of the vision and mission of Cancer Council Queensland (CCQ)

Briefly introduce UNIQUE

- Background and aims
- Recruitment
- Data collection

Discuss Phase 1 of the current study (codesign workshops)

- Participants
- Outcomes

Outline purpose of the interview

Activity 1: Revised recruitment material

Introduce activity

Ask participant to read the study flyer

- Display digital version of the study flyer on the presentation slides

Invite participant to share their feedback on the study flyer

– *e.g., What are your thoughts on the flyer? Would you change anything?*

Ask participant to read the invitation letter

- Display digital version of the invitation letter on the presentation slides

Invite participant to share their feedback on the invitation letter

– *e.g., What are your thoughts on the invitation letter? Would you change anything?*

Activity 2: Survey question

Introduce activity

Explain process for generating and shortlisting the survey questions

Survey question 1: Ask participant to read and respond to the survey question using the chat function (5 minutes)

- Display the survey question on the presentation slides

Introduce the ‘think aloud’ method

- Demonstrate an example

Survey question 1: Ask participant to talk through their interpretation of the survey question using the ‘think aloud’ method

*– e.g., Please read the question out loud and tell me what you’re thinking.*

Ask participant to read other alternative questions and indicate their preference for wording

- Display the survey questions on the presentation slides

Conclusion

Collect demographic data from participant (using structured verbal questions)

Invite participant to share additional thoughts or feedback

Thank participant for their contributions

### **S5 Table.** Responses to the shortlisted and final question in the pretest interviews.

| Question | Participant responses |
| --- | --- |
| Option 1:  Please tell us about your experiences since diagnosis and what would help you most right now? | *Earlier intervention with allied health services whilst undergoing treatment. Personally, I would have benefited from exercise physiology or physio during my treatment to ensure loss of function wasn't so profound when I completed treatment and was supposed to just get back to "normal life". If there were services to help with transport or help with children when you're a primary carer of young children, I wasn't made aware of them. This was the greatest struggle for me as a cancer sufferer – coping with treatment and raising a young family. I was put in touch with a charity who cleaned my house periodically. If CCQ could offer that, it'd be great. Second to that greatest need was obviously financial and if there was financial counselling more readily available that'd also be great.* (P1, cancer survivor, <5 years post-diagnosis) |
|  | *I consider myself one of the lucky cancer survivors. I was diagnosed with prostate cancer [just over a year ago] and underwent a radical prostatectomy [last year]. Tests since then have shown that the surgery has been successful, but the testing will be ongoing every 6 months. So, I have dodged the "bullet" that many other cancer patients go through with ongoing therapies etc. Since diagnosis I received a lot of information from the specialist, prostate nurses at the hospital, my GP, and the local support group. I felt at times there was too much information, too much to take in and too many choices. It was a bit intimidating during an uncertain time. If I was to go through this again, I would probably prefer more discussion type support than the books, flyers, and other papers I received.* (P4, cancer survivor, <5 years post-diagnosis) |
|  | *My mum has recently been diagnosed with breast cancer and I have been caring for her, taking her to the hospital, being as supportive as possible and just present. I feel that this is a very sensitive time for her and sometimes I feel like I am not doing a good job. I think some level of support from someone trained and/or experienced would be ideal to help reduce stress and make her feel normal. I also think she is isolating herself through this process – so assistance in that area would be beneficial.* (P11, previous caregiver, <5 years post-diagnosis) |
|  | *I am the carer for my daughter, who was initially diagnosed with cancer in [3 years ago], had a year of treatment and went into remission, only to be diagnosed with a secondary cancer in [just over 1 year later]. At the time of diagnosis, both times, it was a massive whirlwind, and we had to very quickly organise work schedules, and care for our other children. We were lucky to have family support which made a huge impact, but there was not a significant amount of support available after initial diagnosis to access things like financial planning help, and help advocating to workplaces etc, which would have been invaluable. There was also very little in the way of information we were provided with about what was available. Now we are almost two years post her second diagnosis, there is still a significant impact as a result of the damage caused by the cancers and the treatment. Access to things like counselling for the whole family would be massively beneficial to us now, as would access to assistance with things to support the ongoing physical support needs (for example, access to financial support to help with physiotherapy, etc).* (P13, current caregiver, <5 years post-diagnosis) |
| Option 2:  How has cancer affected your life and what do you need right now? | *Doctors who listen... What I need right now is doctors who actually tell the truth and doctors who listen. That's what I need right now because they're not telling the truth and they're not listening... That's what I need. How cancer affected me? It actually made me go to medical school and read medical books and understand what cancer is, how it actually starts, what the root causes are, and I've actually had to learn how to do this and actually fix it myself. That's how it affected me.* (P3, cancer survivor, unknown diagnosis date) |
|  | *Right now, my family needs stronger bereavement support and post-treatment care. After my five-year-old niece passed way, we received only one follow-up phone call from her oncologist, which left us feeling like just another number in the system. My sister and my brother-in-law are reluctant to ask for help, even though they are struggling. We have had to seek out counselling support through [charity] on our own accord because our social worker at the [hospital] quit her job and moved on suddenly. It would also be helpful to know how we go about re-instigating correspondence with [niece’s] treatment team at the [hospital] in order to ask the questions that we still haven't had answered. For example, how do we go about accessing all of [niece’s] medical files, including her scan results, and her genomic sequencing which was conducted through the Zero program? If CCQ could help answer these questions or provide advice/guidance, that would be extremely helpful.* (P7, previous caregiver, <5 years post-diagnosis) |
|  | *We are based in [small city centre] but since my son diagnosed with LCH [Langerhans cell histiocytosis], we have had to travel to [major city centre] (first stage – every week, now every 3 weeks) that makes things hard as we have another son. It would make things easier if we could get treatment on the Gold Coast. Treatment itself is already stressful but traveling is adding more stress for us.* (P15, current caregiver, <5 years post-diagnosis) |
|  | *Can't work, no income, in incredible pain, family are scared and deeply concerned. Need answers, how and when will treatment start, don’t know what many of these cancer terms and names mean. Will I live, no one seems to know what my cancer is.* (P2, cancer survivor, ≥5 years post-diagnosis) |
| Option 3:  Please tell us how we can support you right now? | *As a cancer patient, it’s hard for me to write this but the support I need is emotional /psychological support – someone to talk to that doesn't judge me for all the horrible thoughts I have and just need to let out. I need someone to help financially because I have no income, [Government welfare] won’t cover me because of my partner’s income, and I have no savings or super options. I am struggling to pay all of my bills, stay financially independent as my own person, and feel that I am not contributing to my family and their needs. I needed physical help to get my daughter to school and attend my appointments, most days I tried to do it myself and often drove myself to appointments when I was too sick, but I was too proud to ask for help. I also guess I figured the support was for people who needed it more than me as they were worse off. I guess I also need help navigating the millions of appointments and information and then help navigating the practical side of recovery if/when I come out on the other side.* (P6, cancer survivor, <5 years post-diagnosis) |
|  | *Assistance with helping my dad get/organise 'regular' GP visits (non-specialist appointments etc), such as transport to and from GP, knowing where to find stock levels for medicines etc (should there be shortages). I have seen some organisations offer free 30-minute discussions with psychologists or crisis counsellors when times get particularly hard or trying. As a carer, being able to get advice on how to lift/support him physically without injuring myself too (or if there are aids or tools I could use too) would be very useful as well.* (P5, current caregiver, <5 years post-diagnosis) |
|  | *Providing support person counselling services. It's about, you know, about how people can get access to support for counselling services for information about how they, as the support person, can be better supporting the person with cancer who’s going through their treatment... Like, you know, just things like getting people to and from treatment. You know, those sorts of things.* (P8, previous caregiver, ≥5 years post-diagnosis) |
|  | *Guidance on what to expect living life with cancer. Possibly more non-formal opportunities to discuss what living with cancer means to work, relationships, kids, finances, and your outlook on life. Access to counselling more promptly and for longer periods.* (P17, cancer survivor, ≥5 years post-diagnosis) |
| Option 4:  What support do you need right now based on your experience? | *Support with financial support, advice, and strategies to cope with everyday living. Emotional support and how I can support my family during this time. Help with logistics and transport to medical appointments and treatment.* (P2, cancer survivor, ≥5 years post-diagnosis) |
|  | *As a female cancer patient, with some difficult diagnostic experiences behind me, I have had more than a few problems with doctors not believing or understanding my symptoms. This happened in the beginning, when I was first diagnosed, and it has continued to happen along the way. Is there any way that CCQ can help me or other female Queenslanders to find the right health help once we are diagnosed or can CCQ assist me with health advocacy? I sometimes feel like there is a gap between me and my doctors. Like I don't have a group or helpful person to assist me with my journey through this awful disease.* (P10, cancer survivor, ≥5 years post-diagnosis) |
| Final question:  Thinking about the past month, how is cancer affecting your life and what do you need? | *This past month has been filled with juggling appointments, rearranging appointments around work commitments. I feel as though I just get life on track for work and my study needs – and my career has something come up that must be addressed straight away. My life is constantly on hold and reactive to my son’s needs. It would be good to be able to plan and actually stick to the plan for a while. It would be good for specialists to understand that sometimes – I can't just drop everything to come from [regional community] to [major city centre]. Life doesn’t work like that. A semi-schedule would be great!* (P16, current caregiver, ≥5 years post-diagnosis) |
|  | *In the past month, cancer has affected my finances as I still am not back at full time work. I have physical issues as well and cannot afford to join a gym etc to help deal with weight gain and loss of strength. It has also affected my ability to do enjoyable things, such as long hikes, bushwalks, so affects my mental health too. Help with financial issues, mental health, and physical exercise programs would be highly beneficial.* (P9, cancer survivor, <5 years post-diagnosis) |
|  | *Thinking about the past month, l feel like cancer takes over a lot of your life, even as a carer. You think about it all the time and want to give your life over to your loved one and other loved ones who are caring. It's difficult to separate your life from theirs and their needs from yours. In terms of what l need, there’s a lot of basic human needs that probably go neglected. For example, the obvious ones like downtime and exercise, but then there are lots of others like having structure and appointments and having some idea of a pathway.* (P12, previous caregiver, ≥5 years post-diagnosis) |
|  | *Physical: pain, vomiting, no appetite, numbness on fingers and toes, emotional swings (sometimes hopeful and sometimes very fearful, scared to have scan or blood results). Financial: may have to reduce working time which will lead to [less] income. Social: hard to tell others, hope to get care and support but scared to get too much empathy. Also, peer support and positive feedback from other patients are very useful.* (P19, cancer survivor, ≥5 years post-diagnosis) |
|  | *I have bowel cancer which has now permanently affected my life both physically, and emotionally. It has left me very immobile due to high doses of radiation and chemo[therapy] treatment. Probably the thing I miss the most, is the support once you have the hospital and it’s this whereby mental health has taken a downturn.* (P20, cancer survivor, <5 years post-diagnosis) |
|  | *I need to use more time to take my mum to hospital. Also, I need to face that she would have more questions [about] her lifespan. That I need to be more positive and show encouragement to her. Mostly just confidence and emotional support are mostly needed. What I would need is someone who have survived or been through similar cancer process could have a group that I would be able to bring her in. So that she can discuss with other patients, and they can also encourage each other. By this way she might be able to get more emotional support.* (P18, current caregiver, <5 years post-diagnosis) |
